# Practical considerations for Ultraviolet-C radiation mediated decontamination of N95 respirator against SARS-CoV-2 virus

**DOI:** 10.1101/2020.11.24.20237917

**Authors:** Guillaume R. Golovkine, Allison W. Roberts, Chase Cooper, Sebastian Riano, Angela M. DiCiccio, Daniel L. Worthington, Jeffrey P. Clarkson, Michael Krames, Jianping Zhang, Ying Gao, Ling Zhou, Scott B. Biering, Sarah A. Stanley

## Abstract

Decontaminating N95 respirators for reuse could mitigate shortages during the COVID-19 pandemic. We tested a portable UV-C light-emitting diode disinfection chamber and found that decontamination efficacy depends on mask model, material and location on the mask. This emphasizes the need for caution when interpreting efficacy data of UV-C decontamination methods.

---

The limited availability of N95 respirators during the SARS-CoV-2 pandemic has forced many healthcare workers to reuse respirators designed for one-time use. The Center for Disease Control (CDC) identified ultraviolet germicidal irradiation (UVGI) as one of 4 most promising methods for N95 decontamination during a crisis capacity situation (*1*).

Although the efficacy of UVGI for decontamination of other viruses, such as influenza, has been investigated (*2–4*), very few studies directly evaluate UV-C mediated inactivation of SARS-CoV-2 on N95 respirators (*5,6)*. Furthermore, most studies are performed using small, flat mask coupons that do not recapitulate angular incidence and shadowing effects caused by the 3D structure of the masks (*7*).

We created a UVGI device for N95 decontamination designed to address these factors via high levels of reflection and enable ease of use via straightforward fixturing and application. The decontamination chamber consists of a metal reflecting box containing high power, commercially available UV LEDs with driver circuitry on metal core printed PCBs mounted on the sidewalls. The LEDs are arrayed in a fashion to optimize exposure dose uniformity across the surface of an N95 respirator and were calibrated to deliver a minimum irradiance of 1 mW/cm^2^ across all locations of the mask (see Appendix).

We tested this device for decontamination of SARS-CoV-2 on two masks models, 3M 1860 and 3M 8210. We analyzed decontamination of 5 different inoculated mask locations (center, top, bottom, right cheek and strap, Figure, panel A). Masks were exposed to UV-C for 0, 300 or 600 seconds. The minimal doses received at each location were greater than 300 and 600 mJ/cm^2^ for the 300 and 600 second exposures, respectively.

**Figure:**
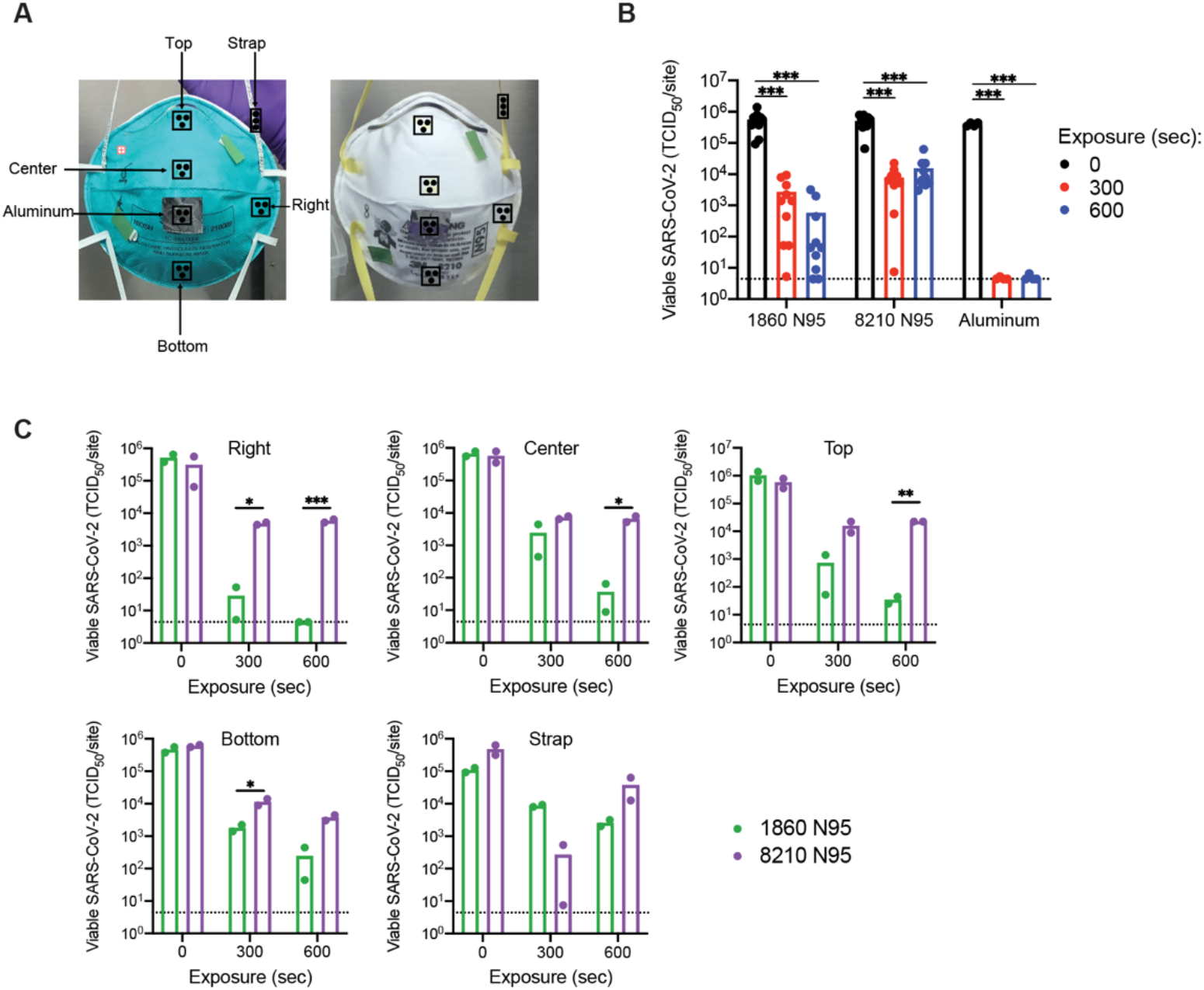
UV-C decontamination of multiple locations on two models of N95 respirators. (A) Schematic of mask inoculation sites. 3M 1860 and 3M 8210 masks were inoculated in five different locations plus an aluminum coupon adhered to the center of each mask. Each site was inoculated with 50ul of 8e^7^ TCID_50_/ml virus, applied as three aliquots of 16.7ul. Inoculated masks were allowed to dry for 3.5 hours at room temperature in a biosafety cabinet before masks were exposed to UV-C irradiation. UV tape was adhered to masks to confirm irradiation. (B and C) Viable SARS-COV-2 recovered from inoculation sites. Viable virus at each inoculation site was quantified by end-point titration on Vero E6 cells and expressed as 50% tissue-culture infectious dose 50 (TCID_50_) per site. Plots show the mean of two replicates from one experiment and are representative of two independent experiments. Dashed lines indicate the limit of detection (LOD), samples with no positive wells are plotted at LOD. (B) Data displayed by mask model. (C) Data displayed by location of inoculation. ∗ p < 0.05, ∗∗ p < 0.01, ∗∗∗ p < 0.001.

Industry standards consider 3 log_10_ reductions as effective for decontamination. Both UV-C doses achieved a 5 log_10_ reduction in virus on the aluminum control coupons (Figure, panel B and Table), validating the efficacy of our UV-C device to eliminate SARS-CoV-2 on non-porous material. However, the 300 second exposure was insufficient for decontamination when averaging locations across the masks (Figure, panel B and Table). The 600 second exposure effectively decontaminated the 3M 1860 masks but failed to decontaminate 3M 8210 masks (Figure, panel B and Table). Notably, there was little difference between the 300 and 600 second doses on the 8210 masks regardless of location, suggesting that increased exposure time does not achieve higher levels of decontamination of this mask surface

**Table:** Average log_10_ reduction in viable SARS-CoV-2 recovered

|  | 300 sec |  | 600 sec |  |
| --- | --- | --- | --- | --- |
|  | 3M 1860 | 3M 8210 | 3M 1860 | 3M 8210 |
| Total mask | 2.67 | 2.05 | 3.74 | 1.68 |
| Aluminum | 4.95 | 4.92 | 4.95 | 4.86 |
| Right | 4.25 | 1.81 | 5.06 | 1.73 |
| Center | 2.44 | 1.90 | 4.26 | 1.94 |
| Top | 3.15 | 1.56 | 4.47 | 1.41 |
| Bottom | 2.41 | 1.72 | 3.28 | 2.21 |
| Strap | 1.10 | 3.24 | 1.63 | 1.10 |

While the reduction averaged across the entire mask was greater than 3 log_10_ for the 3M 1860 mask at 600 seconds, there were important variations between different locations on the mask (Figure, panel C and Table). Irradiation doses were calculated using a representative N95 mask with integrated irradiance sensors (Appendix). We determined that the smaller reduction in viral titer at the bottom location correlates with a lower irradiation dose received at this location. Conversely, the Right location received the highest irradiation dose and demonstrated effective decontamination of the 3M 1860 masks at the 300 sec and 600 sec exposures. However, neither dose at the Right location was sufficient for decontamination of the 3M 8210 masks (Figure, panel C), despite receiving the largest dose. Furthermore, the straps were difficult to decontaminate, with large variability (Figure, panel C), a result observed for other viruses (4). We hypothesize that this is due to the strap material and potential shadowing effects caused by twists in the strap during exposure to UV-C.

These results suggest that mask material is a major factor in the ability of UV-C to decontaminate an N95 respirator, and that the 1860 facepiece (hydrophobic polypropylene shell) is more appropriate for UV-C decontamination than the 1860 strap (braided polyisoprene), the 8210 facepiece (polyester), or the 8210 straps (thermoplastic elastomers).

While UV-C is an attractive method for decontamination of PPE when applied at appropriate doses that do not compromise material integrity and device functionality, our findings suggest that efficacy for individual mask models should be evaluated for a given UV-C device. Our results as well as the recent study by Ozog et al. (*6*) indicate that while the facepieces of some mask models can be successfully decontaminated using UV-C, others are incompatible with this method of SARS-CoV-2 decontamination. Important factors to consider are the 3D structure of the mask and corresponding differences in irradiation dose received in some mask locations which can significantly influence the efficacy of decontamination. The straps may be particularly difficult to decontaminate and may require the use of a secondary method of decontamination for the straps in addition to UVGI (*8*). However, UVC LED technology is improving rapidly and future devices will offer higher irradiation levels, improving penetration of UVGI and/or shortening exposure times. The identification of existing N95 models that are most suited for UV-C based decontamination or the creation of new mask models for this purpose would be important milestones that could help mitigate future N95 shortages.

## Data Availability

All data is included in the manuscript as individual data points on graphs.

## Appendix Text

### UV-C dose measurements

The UVC LEDs were commercially available products from Bolb, Inc.: Surface Mount Type SMD6060, with peak emission wavelength of 272 nm, full-width-at-half-maximum of 9.5 nm, and outputs of 100 mW (250 mA) or 140 mW (350 mA) per LED. Website: www.bolb.co. The LED emission is narrow, at a wavelength of 272 nm. The range of irradiance incidents upon the surface on 3M 1860 respirators was 1.5 to 3.0 mW/cm^2^. To account for non-uniformity across the surface of a respirator, the irradiance at each inoculation site was measured using a custom N95 respirator with calibrated sensors (Appendix Figure 1).

### SARS-CoV-2 decontamination studies

#### Virus preparation and stock titration

The SARS-CoV-2 strain used was USA-WA1/2020. Viral stocks were obtained from the Biodefense and Emerging Infections Research Resources Repository. BEI stocks were amplified in Vero-E6 cells to produce virus passage 1 and passaged again in Calu-3 cells to produce virus passage 2 which was used for experiments. In brief, for passage 1, 50 ul of the BEI stock was inoculated onto T-175 flasks of Vero-E6 cells and allowed to propagate until 50% cytopathic effect (CPE) was achieved (∼48 hours post infection) at which time flasks were lysed through 1 round of freeze and thaw, then supernatants were collected and clarified through a gentle spin step (1500 rpm for 5 mins). The clarified viral supernatants were aliquoted and frozen down at −80C. Aliquots were thawed for titration or production of virus passage 2 which was done the same as above except using Calu-3 human lung epithelial cells. Viral stocks were propagated in Calu-3 cells grown in Dulbecco’s Modified Eagle Medium (DMEM) containing 10% FBS and penicillin/streptomycin. The concentration of viral stocks was assessed by TCID_50_ assay using Vero-E6 cells and was determined to be 8 x 10^7^ TCID_50_/ml.

#### Mask inoculation

All mask locations (center, top, bottom, right, aluminum coupon and strap) were inoculated with 3 aliquots of 16.67ul, for a total of 50ul, of virus stock. Masks were left to dry for 3.5 hours in a biosafety cabinet. Straps of 3M 1860 masks were inoculated with 50ul of SARS-CoV-2 only 10 minutes before irradiation because optimization experiments showed that virus viability on this material decreased with excessive drying (unpublished data). Desiccation on mask facepieces for 3.5 hours did not significantly affect virus concentration (Appendix Figure 2).

#### Ultraviolet Light

Immediately before irradiation, 2 pieces of UV tape were added to the upper right and lower left corners of the mask. Masks are placed into the device by attaching the straps on mounting points located at the top and bottom. Masks were irradiated for 300 sec or 600 sec. A picture of the mask was taken after irradiation to document UV tape change of color.

#### Virus titration

Inoculated regions of the mask were cut out using 12mm biopsy punches. Mask punches, strap pieces and aluminum coupons were incubated in 1.4ml (mask punches and aluminum coupons) or 2ml (strap pieces) of DMEM (Sigma-Aldrich) supplemented with 10% fetal bovine serum, 100 U/mL penicillin, and 100 μg/mL streptomycin for a minimum of 30 minutes. Virus was quantified by TCID_50_ assay by incubating Vero E6 cells in 96 well plates with 10-fold serial dilutions in 8-fold of incubation media. Five days after inoculation, cytopathic effect was scored visually, defined as any virus induced cell death or change in cell morphology and the TCID_50_ was calculated.

#### Appendix Table

**Appendix Table:** calculated doses delivered at each location for each exposure

|  | 300 sec | 600 sec |
| --- | --- | --- |
| Right | 810 | 1620 |
| Center | 570 | 1140 |
| Top | 450 | 900 |
| Bottom | 450 | 900 |
| Strap | N/A | N/A |

Doses are calculated based on irradiance measurements made with the custom N95 respirator with calibrated sensors. Units are in mJ/cm^2^.

**Appendix Figure 1:**
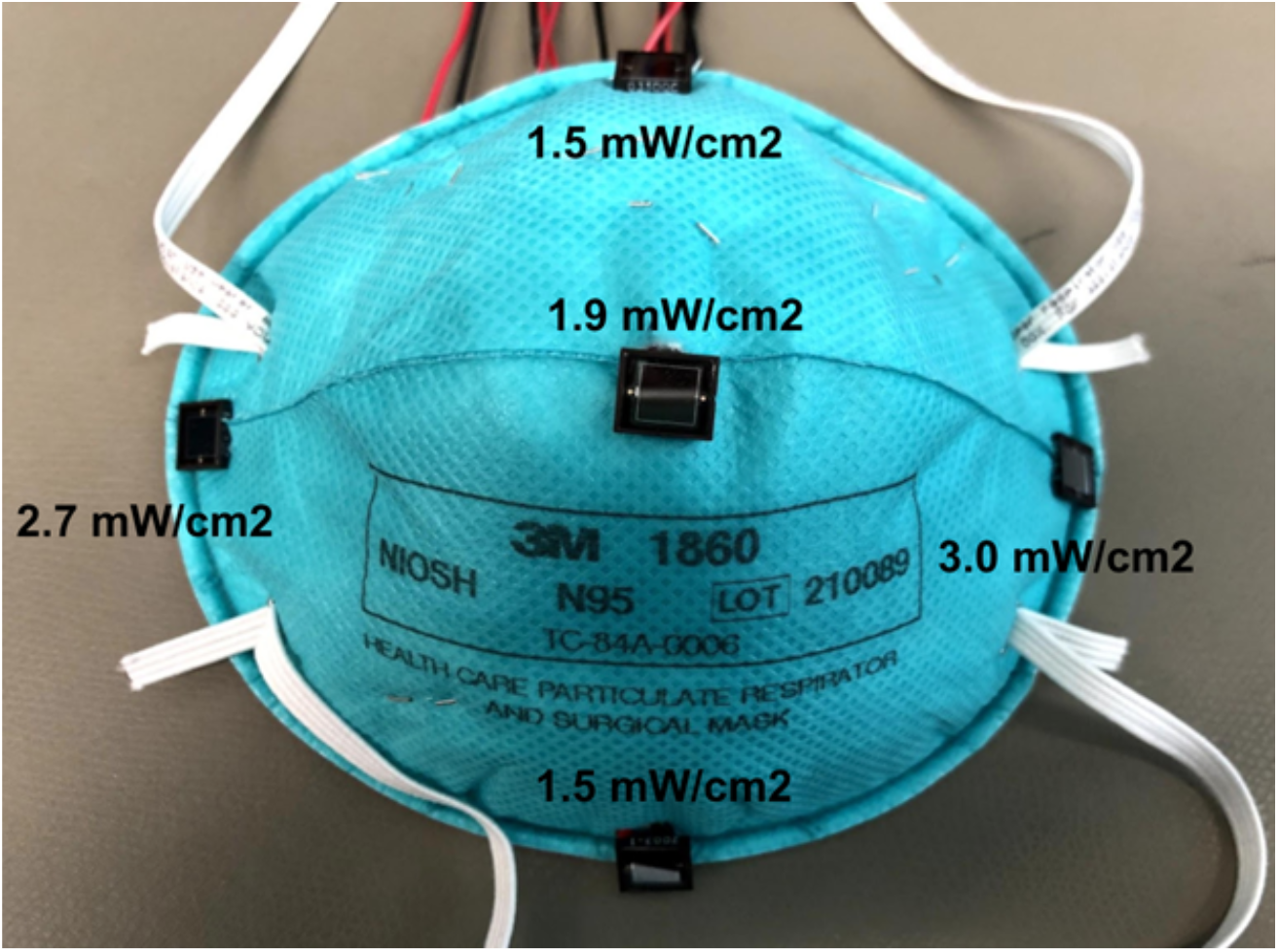
custom N95 respirator with calibrated sensors used for the measurement and according irradiance measurement at each site.

**Appendix Figure 2:**
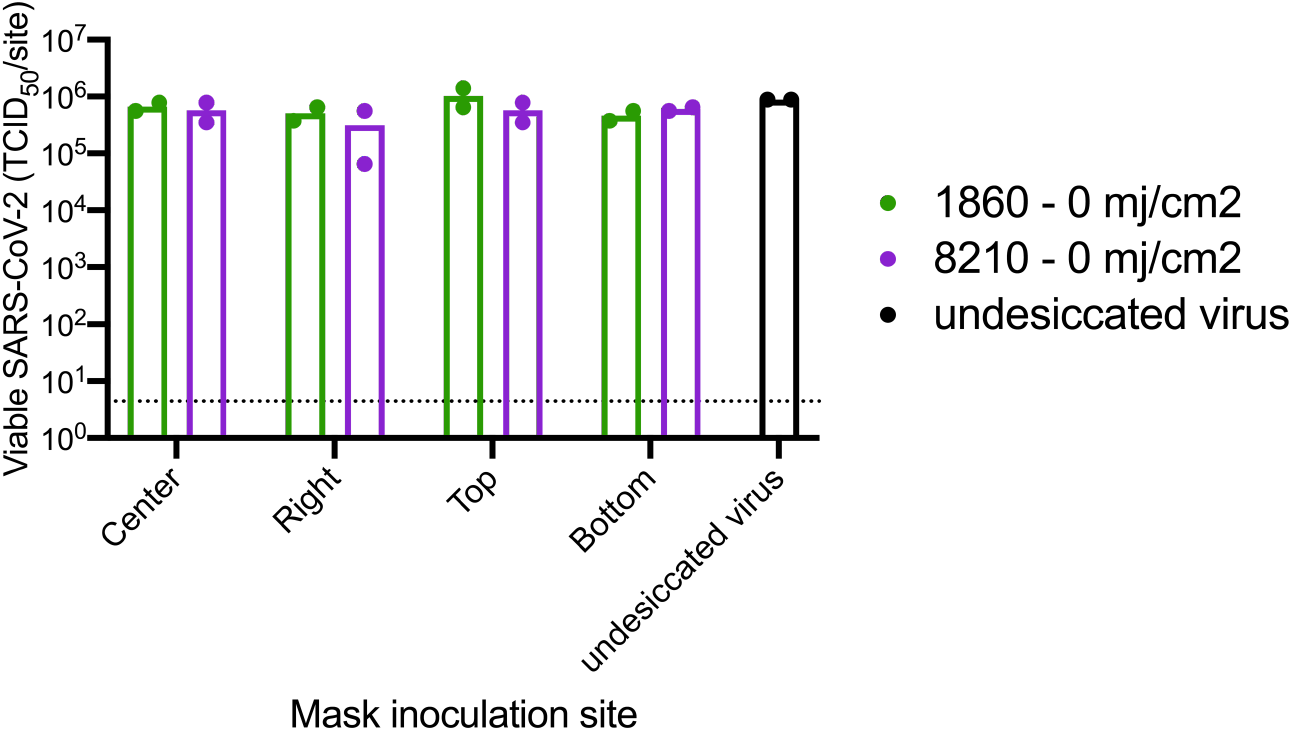
comparison of virus recovery from unirradiated mask sites after virus desiccation to control virus added directly into recovery media.

## Acknowledgments

We thank Verily employees Greg Arcenio, Beth Bosworth, Warren Cai, Mike Chen, Junjia Ding, Tim English, Chopin Hua, David Heinz, Kyle Nichols, Supriyo Sinha for their valuable contributions and feedback.

## Disclaimers

All Verily authors and contributors were full time employees of Verily Life Sciences during their respective contributions to this effort. No financial compensation was received outside of the contributors’ regular monetary and stock compensation due to their employment at Verily Life Sciences.

